# HIV infection and COVID-19 death: population-based cohort analysis of UK primary care data and linked national death registrations within the OpenSAFELY platform

**DOI:** 10.1101/2020.08.07.20169490

**Authors:** K Bhaskaran, CT Rentsch, B MacKenna, A Schultz, A Mehrkar, C Bates, RM Eggo, CE Morton, S Bacon, P Inglesby, IJ Douglas, AJ Walker, HI McDonald, J Cockburn, EJ Williamson, D Evans, HJ Forbes, HJ Curtis, W Hulme, J Parry, F Hester, S Harper, SJW Evans, L Smeeth, B Goldacre

## Abstract

**Background:** It is unclear whether HIV infection is associated with risk of COVID-19 death. We aimed to investigate this in a large-scale population-based study in England.

**Methods:** Working on behalf of NHS England, we used the OpenSAFELY platform to analyse routinely collected electronic primary care data linked to national death registrations. People with a primary care record for HIV infection were compared to people without HIV. COVID-19 death was defined by ICD-10 codes U07.1 or U07.2 anywhere on the death certificate. Cox regression models were used to estimate the association between HIV infection and COVID-19 death, initially adjusted for age and sex, then adding adjustment for index of multiple deprivation and ethnicity, and finally for a broad range of comorbidities. Interaction terms were added to assess effect modification by age, sex, ethnicity, comorbidities and calendar time.

**Results:** 17.3 million adults were included, of whom 27,480 (0.16%) had HIV recorded. People living with HIV were more likely to be male, of black ethnicity, and from a more deprived geographical area than the general population. There were 14,882 COVID-19 deaths during the study period, with 25 among people with HIV. People living with HIV had nearly three-fold higher risk of COVID-19 death than those without HIV after adjusting for age and sex (HR=2.90, 95% CI 1.96-4.30). The association was attenuated but risk remained substantially raised, after adjustment for deprivation and ethnicity (adjusted HR=2.52, 1.70-3.73) and further adjustment for comorbidities (HR=2.30, 1.55-3.41). There was some evidence that the association was larger among people of black ethnicity (HR = 3.80, 2.15-6.74, compared to 1.64, 0.92-2.90 in non-black individuals, p-interaction=0.045)

**Interpretation:** HIV infection was associated with a markedly raised risk of COVID-19 death in a country with high levels of antiretroviral therapy coverage and viral suppression; the association was larger in people of black ethnicity.

## Introduction

Since it emerged in late 2019, SARS-CoV-2, the virus that causes coronavirus disease-19 (COVID-19), has infected over 14 million people worldwide, causing over 600,000 deaths to date.^1^ Older age and male gender have been strongly associated with more severe outcomes; several comorbidities, including those that involve immunosuppression, also appear to be associated with higher risk of COVID-19 death.^2^ However, there has been limited evidence to date on how human immunodeficiency virus (HIV) infection may affect risk of poor outcomes from COVID-19.^3^

There is mixed evidence on the importance of HIV to previous respiratory virus epidemics. HIV has been associated with a higher risk of severe outcomes from respiratory infections including seasonal influenza,^4,5^ and people living with HIV at any stage of infection are considered a clinical risk group in seasonal influenza vaccination guidance in the UK.^6^ But the importance of HIV infection to outcomes from 2009 H1N1 pandemic influenza was unclear, with no substantive evidence that HIV-infected individuals were at higher risk of being infected or experience poor outcomes, unless at an advanced stage of immunosuppression.^7^ Evidence from the present SARS-CoV-2 pandemic is limited. A large population-based cohort study in South Africa found COVID-19 mortality risk among people living with HIV to be double the risk of those without HIV.^8^ A high prevalence of critical illness was observed among HIV-infected patients with COVID-19 in Madrid, though there was no non-HIV comparison group.^9^ Other small studies of hospitalised patients have failed to detect any raised risk of severe outcomes in people living with HIV.^10,11^

We therefore aimed to investigate the association between HIV infection and COVID-19 death using population-based data from England. We conducted our analysis within the OpenSAFELY platform which includes large-scale primary care data from 40% of the English population linked to national death registrations. Information on antiretroviral treatment and viral load was not available for this study, due to the health service delivering these treatments through specialist clinics, and restrictions on the sharing of data that are deemed potentially sensitive.^12^ However, in the UK, 94% of individuals are treated with antiretroviral drugs and have undetectable viral load,^13^ thus our findings should generalise to settings with similarly high levels of HIV control, and may represent a “best-case” scenario for associations between HIV infection and COVID-19 outcomes in other settings.

## Methods

### Study design and study population

A retrospective cohort study comparing the risk of COVID-19 death among people living with and without HIV was carried out within OpenSAFELY, a new data analytics platform in England created to address urgent COVID-19 related questions, which has been described previously.^2^ We used routinely-collected electronic data from primary care practices using The Phoenix Partnership (TPP) SystmOne software, covering approximately 40% of the population in England, linked to Office of National Statistics (ONS) death registrations. We included all adults (aged 18 years or over) alive and under follow-up on 1^st^ February 2020, and with at least one year of continuous GP registration prior to this date, to ensure that baseline data could be adequately captured. We excluded people with missing age, sex, or index of multiple deprivation, since these are likely to indicate poor data quality.

### Outcome, exposure and covariates

The outcome was COVID-19 death, defined as a record for death in linked ONS data with the ICD-10 codes U07.1 (“COVID-19, virus identified”) or U07.2 (“COVID-19, virus not identified”) anywhere on the death certificate.^14^

The main exposure was HIV status, and covariates considered in the analysis included age (at 1^st^ February 2020, grouped as 18-39, 40-49, 50-59, 60-69, 70-79 and ≥80 years for descriptive analysis, and parametrised as a 4-knot restricted cubic spline in regression models), sex, ethnicity (White, Mixed, South Asian, Black, Other, based on self-report using categories from the UK census), obesity (categorised as class I [body mass index 30-34.9kg/m^2^], II [35-39.9kg/m^2^], III [≥40kg/m^2^]), smoking status (never, former, current), index of multiple deprivation quintile (derived from the patient’s postcode at lower super output area level), and comorbidities considered potential risk factors for severe COVID-19 outcomes, namely diagnosed hypertension, chronic respiratory diseases other than asthma, asthma (categorised by use of oral steroids), chronic heart disease, diabetes (categorised according to the most recent glycated haemoglobin (HbA1c) recorded in the 15 months prior to 1^st^ February 2020), non-haematological and haematological cancer (both categorised by recency of diagnosis, <1, 1-4.9, ≥5 years), reduced kidney function (categorised by estimated glomerular filtration rate derived from the most recent serum creatinine measure (30-<60, <30 mL/min/1.73m^2^), chronic liver disease, stroke or dementia, other neurological disease (motor neurone disease, myasthenia gravis, multiple sclerosis, Parkinson’s disease, cerebral palsy, quadriplegia or hemiplegia, and progressive cerebellar disease), organ transplant, asplenia (splenectomy or a spleen dysfunction, including sickle cell disease), rheumatoid arthritis/lupus/psoriasis, and other immunosuppressive conditions (permanent immunodeficiency ever diagnosed, or aplastic anaemia or temporary immunodeficiency recorded within the last year).

Information on HIV and all covariates was obtained by searching TPP SystmOne records prior to 1^st^ February 2020 for specific coded data, based on a GP subset of SNOMED-CT mapped to Read version 3 codes. Over 70 codes for HIV were included, but >90% of people living with HIV were identified from just four codes, namely: “43C3. – HIV positive” (73%), “X70M6 – HIV infection” (10%), “Xa0ye – HIV” (4%), and “X80bg – HIV antibody” (4%). All codelists, along with detailed information on their compilation is available at https://codelists.opensafely.org for inspection and re-use by the wider research community.

### Statistical analysis

Follow-up time for COVID-19 mortality was from 1^st^ February 2020 until the date of COVID-19 death or 22^nd^ June 2020, which was the last date for which mortality data were complete. The competing risk of death from causes other than COVID-19 was accounted for by censoring non-COVID deaths, so our analysis focussed on cause-specific hazards.^15^ Crude rates of COVID-19 in the HIV and non-HIV groups were calculated, and we then used Cox regression models to estimate the association between HIV infection and COVID-19 mortality, initially adjusted for age and sex; then adding adjustment for index of multiple deprivation and ethnicity; and finally adding adjustment for all comorbidities listed above to minimise confounding; it should be noted that the fully adjusted model would not capture any effect of HIV mediated through an increased risk of the included comorbidities. We additionally adjusted for history of hepatitis C infection in a post-hoc analysis, as this might be a marker of injection drug use. Models were stratified by the Sustainability and Transformation Partnership (STP, a UK National Health Service administrative region) of the patient’s general practice to allow for geographical differences in baseline hazards. Multiple imputation (10 imputations) was used to account for missing ethnicity, with the imputation model including all covariates from the main model and an indicator for the outcome. Those with missing body mass index were assumed to be non-obese, and those with missing smoking data were assumed to be never-smokers; we did not use multiple imputation for these variables as they are expected to be missing not at random in UK primary care.^16^ In sensitivity analyses, we excluded individuals with missing data (complete case analysis). Proportional hazards were checked by: (i) by fitting an interaction between analysis time and the HIV variable, with analysis time categorised as 0-60, 60-90, ≥90 days from 1^st^ February 2020, chosen to respectively capture the period before social distancing policies in the UK would have impacted mortality, the period of peak COVID-19 mortality, and the period during which restrictions began to be eased; and (ii) by testing the slope of the Schoenfeld residuals (among those with complete ethnicity only).

To investigate effect modification by variables considered *a priori* to be of key potential importance, we fitted interaction terms (one at a time) between HIV status and age (<60, ≥60 years), sex, ethnicity (black vs other), and presence of comorbidities (any versus none of the comorbidities included in the analysis – a sensitivity analysis excluded hypertension from this list since a past code for hypertension may not reflect any ongoing clinical disease). These variables were dichotomised due to limited power. P-values (two-sided) were from Wald tests on the interaction terms.

Cumulative mortality curves, standardised to adjust for different covariate distributions in the HIV and non-HIV groups, were generated. First, a Royston-Parmar model with the same covariates as the fully adjusted Cox model was fitted, with the baseline hazard modelled using a 3 degrees-of-freedom spline.^17^ The survival function was predicted from this model for every individual with HIV, and averaged to produce the curve for the HIV group. To produce the standardised comparison curve, the survival functions were predicted and averaged again for the same individuals, but with HIV status set to 0.

### Information Governance and Ethics

NHS England is the data controller; TPP is the data processor; and the key researchers on OpenSAFELY are acting on behalf of NHS England. OpenSAFELY is hosted within the TPP environment which is accredited to the ISO 27001 information security standard and is NHS IG Toolkit compliant;^18,19^ patient data are pseudonymised for analysis and linkage using industry standard cryptographic hashing techniques; all pseudonymised datasets transmitted for linkage onto OpenSAFELY are encrypted; access to the platform is via a virtual private network (VPN) connection, restricted to a small group of researchers who hold contracts with NHS England and only access the platform to initiate database queries and statistical models. All database activity is logged; only aggregate statistical outputs leave the platform environment following best practice for anonymisation of results such as statistical disclosure control for low cell counts.^20^ The OpenSAFELY platform adheres to the data protection principles of the UK Data Protection Act 2018 and the EU General Data Protection Regulation (GDPR) 2016. In March 2020, the Secretary of State for Health and Social Care used powers under the UK Health Service (Control of Patient Information) Regulations 2002 (COPI) to require organisations to process confidential patient information for the purposes of protecting public health, providing healthcare services to the public and monitoring and managing the COVID-19 outbreak and incidents of exposure.^21^ Taken together, these provide the legal bases to link patient datasets on the OpenSAFELY platform. This study was approved by the Health Research Authority (REC reference 20/LO/0651) and by the LSHTM Ethics Board (ref 21863).

## Results

27,480 adults living with HIV and 17,255,425 without HIV were included (Figure 1). People with HIV had similar median age, but a narrower age distribution overall, compared to those without HIV (Table 1). People living with HIV were more likely to be male, of black ethnicity, and from a more deprived area. 921 (3.4%) individuals with HIV had chronic liver disease, compared to 99,214 (0.6%) of those in the comparison group. 1,491 individuals with HIV (5.4%) had a history of hepatitis C compared to 38,235 (0.2%) of those without HIV.

**Figure 1:**
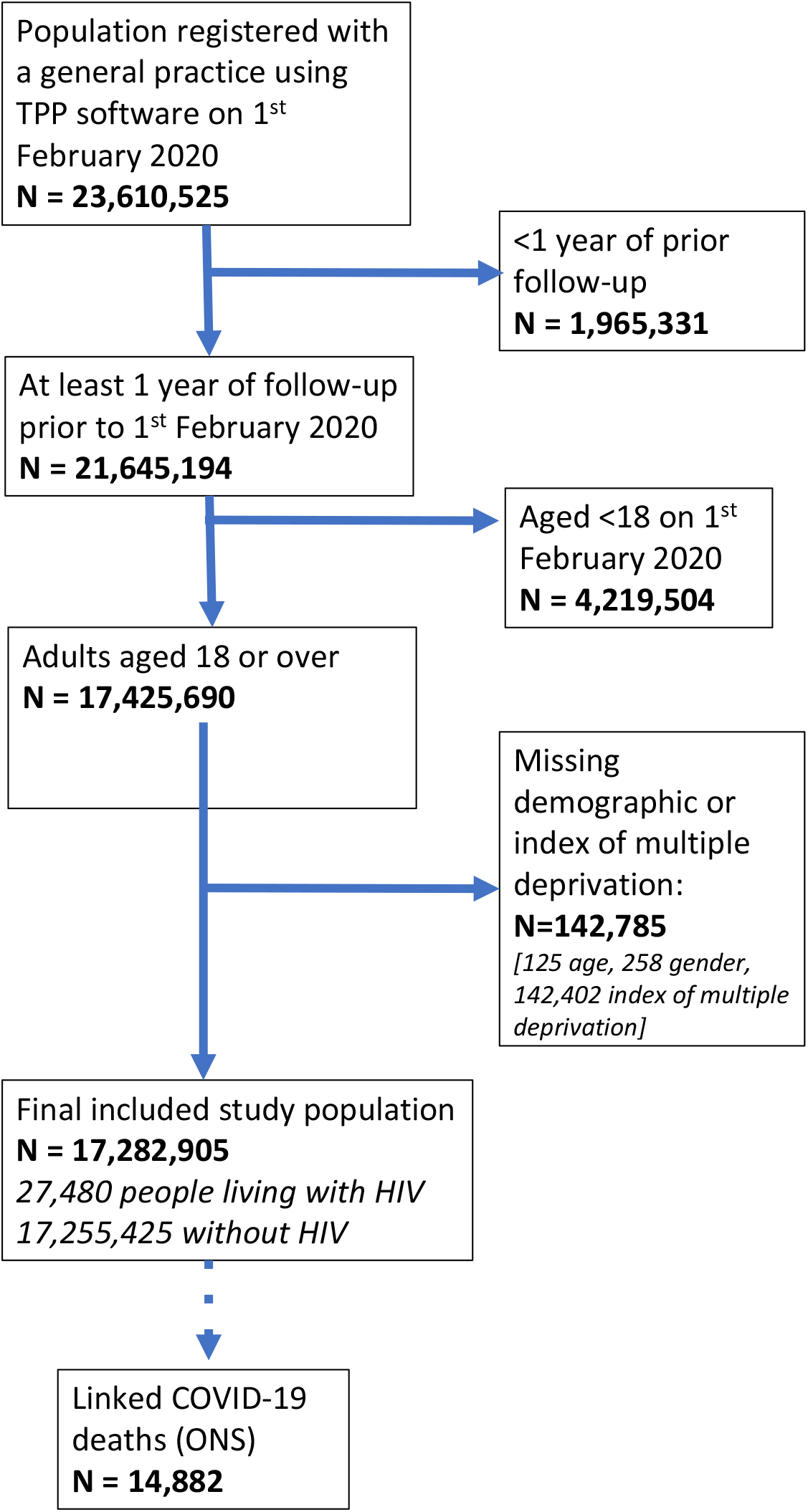
Flow chart of study participants.

**Table 1:** Demographic characteristics of people with and without HIV in OpenSAFELY.

| (cells are n (%) unless stated) |  | HIV group | No HIV group |
| --- | --- | --- | --- |
| N |  | 27,480 (100.0) | 17,255,425 (100.0) |
| Age (yrs) | 18-39 | 6,625 (24.1) | 5,908,794 (34.2) |
|  | 40-49 | 8,486 (30.9) | 2,842,155 (16.5) |
|  | 50-59 | 8,093 (29.5) | 3,043,868 (17.6) |
|  | 60-69 | 3,130 (11.4) | 2,390,044 (13.9) |
|  | 70-79 | 937 (3.4) | 1,938,734 (11.2) |
|  | 80+ | 209 (0.8) | 1,131,830 (6.6) |
|  | Median (IQR) | 48 (40-55) | 49 (34-64) |
| Sex | Male | 17,780 (64.7) | 8,614,886 (49.9) |
|  | Female | 9,700 (35.3) | 8,640,539 (50.1) |
| Body mass index (kg/m<sup>2</sup>)* | Not obese | 21,539 (78.4) | 13,488,390 (78.2) |
|  | 30-34.9 (Obese class I) | 3,777 (13.7) | 2,381,893 (13.8) |
|  | 35-39.9 (Obese class II) | 1,475 (5.4) | 921,864 (5.3) |
|  | ≥40 (Obese class III) | 689 (2.5) | 463,278 (2.7) |
| Smoking status* | Never | 13,398 (48.8) | 8,634,105 (50.0) |
|  | Former | 7,922 (28.8) | 5,685,069 (32.9) |
|  | Current | 6,160 (22.4) | 2,936,251 (17.0) |
| Ethnicity | White | 12,708 (46.2) | 10,859,301 (62.9) |
|  | Mixed | 1,390 (5.1) | 168,649 (1.0) |
|  | South Asian | 1,175 (4.3) | 1021,252 (5.9) |
|  | Black | 7,143 (26.0) | 332,971 (1.9) |
|  | Other | 645 (2.3) | 319,845 (1.9) |
|  | Missing | 4,419 (16.1) | 4,553,407 (26.4) |
| Index of Multiple Deprivation | 1 (least deprived) | 2,790 (10.2) | 3,494,398 (20.3) |
|  | 2 | 4,127 (15.0) | 3,473,416 (20.1) |
|  | 3 | 5,108 (18.6) | 3,480,657 (20.2) |
|  | 4 | 6,863 (25.0) | 3,475,019 (20.1) |
|  | 5 (most deprived) | 8,592 (31.3) | 3,331,935 (19.3) |
| **Comorbidities** |  |  |  |
| Hypertension |  | 5,290 (19.3) | 3,666,002 (21.2) |
| Chronic respiratory disease |  | 1,095 (4.0) | 703,335 (4.1) |
| Asthma | With no oral steroid use | 3,460 (12.6) | 2,454,236 (14.2) |
|  | With oral steroid use | 433 (1.6) | 291,606 (1.7) |
| Chronic heart disease |  | 1,444 (5.3) | 1,167,008 (6.8) |
| Diabetes | With HbA1c<58 mmol/mol | 1,521 (5.5) | 1,037,513 (6.0) |
|  | With HbA1c≥58 mmol/mol | 739 (2.7) | 485,895 (2.8) |
|  | With no recent HbA1c measure | 449 (1.6) | 193,665 (1.1) |
| Cancer (non-haematological) | Diagnosed < 1 year ago | 108 (0.4) | 80,004 (0.5) |
|  | Diagnosed 1-4.9 years ago | 372 (1.4) | 233,977 (1.4) |
|  | Diagnosed ≥5 years ago | 804 (2.9) | 541,921 (3.1) |
| Haematological malignancy | Diagnosed < 1 year ago | 30 (0.1) | 8,691 (0.1) |
|  | Diagnosed 1-4.9 years ago | 114 (0.4) | 27,648 (0.2) |
|  | Diagnosed ≥5 years ago | 421 (1.5) | 63,076 (0.4) |
| Reduced kidney function |  |  |  |
|  | Estimated GFR 30-60 | 1,427 (5.2) | 1,006,298 (5.8) |
|  | Estimated GFR <30 | 134 (0.5) | 77,964 (0.5) |
| Chronic liver disease |  | 921 (3.4) | 99,214 (0.6) |
| Stroke or dementia |  | 559 (2.0) | 389,828 (2.3) |
| Other neurological disease |  | 239 (0.9) | 170,336 (1.0) |
| Organ transplant |  | 72 (0.3) | 19,933 (0.1) |
| Asplenia |  | 89 (0.3) | 27,845 (0.2) |
| Rheumatoid arthritis/lupus/psoriasis |  | 1,233 (4.5) | 877,666 (5.1) |
| Other immunosuppressive conditions |  | 53 (0.2) | 4,211 (0.0) |

\*missing BMI included in 'not obese' (HIV group: 4,550, non-HIV group: 3,737,207; missing smoking included in 'never smoker' (HIV group: 4,550, non-HIV group: 3,737,207). N with any comorbidity (of those listed) = 12,984 (47.3%) in HIV group and 7,914,272 (45.9%) in non-HIV group; N with ever hepatitis C = 1,491 (5.4%) in HIV group and 38,235 (0.2%) in non-HIV group.

There were 25 COVID-19 deaths among people with HIV, and 14,857 among those without HIV (Table 2, Table 3). Estimated cumulative mortality, standardised to the covariate distribution of the HIV group, was 0.087% (95% CI 0.056-0.134) and 0.038% (0.036-0.040) in people with and without HIV respectively, at the last date of mortality data coverage (22^nd^ June, Figure 2).

**Table 2:** Characteristics of people living with HIV who died with COVID-19 as a listed cause of death.

|  | N (%) |
| --- | --- |
| **Overall** | 25 |
| **Age in years** |  |
| <60 | 10 (40) |
| ≥60 | 15 (60) |
| **Sex** |  |
| Male | 18 (72) |
| Female | 7 (28) |
| **Body mass index** |  |
| Not obese | 15 (60) |
| Obese | 10 (40) |
| **Ethnicity** |  |
| Black<sup>+</sup> | 11 (44) |
| White/other/unknown | 14 (56) |
| **Common comorbidities\*** |  |
| Hypertension | 15 (60) |
| Diabetes | 14 (56) |
| Reduced kidney function** | 9 (36) |
| Any comorbidity*** [excluding hypertension] | 23 (84) [21 (92)] |

<sup>+</sup> self-report as African, Caribbean or other Black
\*all other individual comorbidities were present in ≤5 individuals and redacted as per our protocol.
\*\*estimated glomerular filtration rate (eGFR) <30 ml min<sup>-1</sup> per 1.73m<sup>2</sup>
\*\*\* from those listed in Table 1

**Table 3:** COVID-19 mortality among patients with and without HIV.

|  |  |  |
| --- | --- | --- |
| **HIV group** |  |  |
| COVID-19 deaths / 1000 person-years | 25 / 10.68 |  |
| Rate per 1000 person-years (95% CI) | 2.34 (1.58-3.47) |  |
| **No HIV group** |  |  |
| COVID-19 deaths / 1000 person-years | 14857 / 6696 |  |
| Rate per 1000 person-years (95% CI) | 2.22 (2.18-2.25) |  |
| **Hazard ratio for COVID-19 death\*** |  |  |
| **(HIV versus no HIV)** |  |  |
| Adjusted for age-sex | 2.90 (1.96-4.30) |  |
| + index of multiple deprivation, ethnicity | 2.52 (1.70-3.73) |  |
| + all covariates<sup>+</sup> | 2.30 (1.55-3.41) |  |
| *Stratified by age* |  |  |
| <60 years | 2.86 (1.53-5.33) | *(p-interaction 0.407)* |
| ≥60 years | 2.03 (1.22-3.38) |  |
| *Stratified by sex* |  |  |
| Male | 2.78 (1.32-5.86) | *(p-interaction 0.564)* |
| Female | 2.15 (1.35-3.42) |  |
| *Stratified by ethnicity<sup>&</sup>* |  |  |
| Black | 3.80 (2.15-6.74) | *(p-interaction 0.045)* |
| White/South Asian/Other | 1.64 (0.92-2.90) |  |
| *Stratified by comorbidity status<sup>~</sup>* |  |  |
| No recorded comorbidities | 1.02 (0.26-4.10) | *(p-interaction 0.212)* |
| At least one comorbidity | 2.57 (1.71-3.88) |  |
| *Stratified by days since 1<sup>st</sup> February 2020* |  |  |
| 0-59 | 4.05 (1.81-9.05) | *(p-interaction 0.258)* |
| 60-89 | 1.79 (1.01-3.15) |  |
| 90+ | 1.15 (0.65-2.03) |  |

\*models including ethnicity used multiple imputation to handle missing data; <sup>+</sup>adjusted for age, sex, obesity, smoking, ethnicity, index of multiple deprivation, high blood pressure or diagnosed hypertension, chronic respiratory disease, asthma, chronic cardiac disease, diabetes, non-haematological cancer, haematological cancer, chronic liver disease, stroke/dementia, other neurological disease, reduced kidney function, organ transplant, asplenia, rheumatoid arthritis/lupus/psoriasis, other immunosuppressive conditions; <sup>&</sup>a similar pattern was seen in a direct comparison between black (HR=3.80, 2.16-6.74) and white (HR=1.81, 0.94-3.48) among those with complete ethnicity data. <sup>~</sup>based on the comorbidities listed in Table 1 – excluding hypertension from the list of comorbidities gave stratified HRs of 1.57 (0.59-4.19) and 2.52 (1.64-3.87) for those without and with comorbidities respectively.

**Figure 2:**
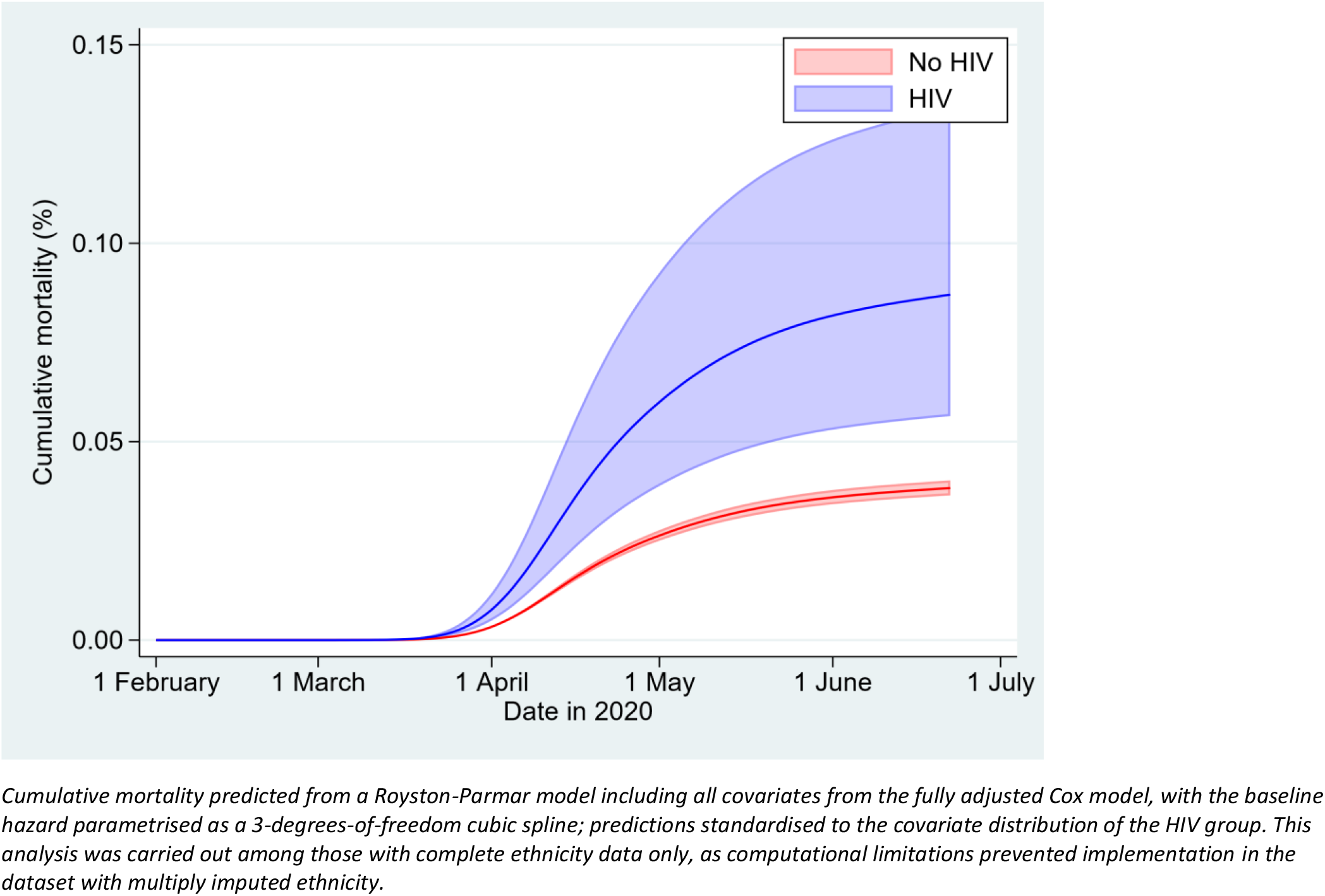
Cumulative mortality by HIV status with 95% CI, standardised to covariate distribution of the HIV group.

HIV was associated with a nearly 3-fold higher risk of COVID-19 death adjusting for age and sex (HR=2.90, 95% CI 1.96-4.30, Table 3 and Figure 3). This attenuated slightly after further adjustment for index of multiple deprivation and ethnicity (HR=2.52, 1.70-3.73), and then obesity, smoking, and comorbidities (HR=2.30, 1.55-3.41). Additionally adjusting for hepatitis C in a post-hoc analysis made little difference (HR=2.28, 1.53-3.38).

**Figure 3:**
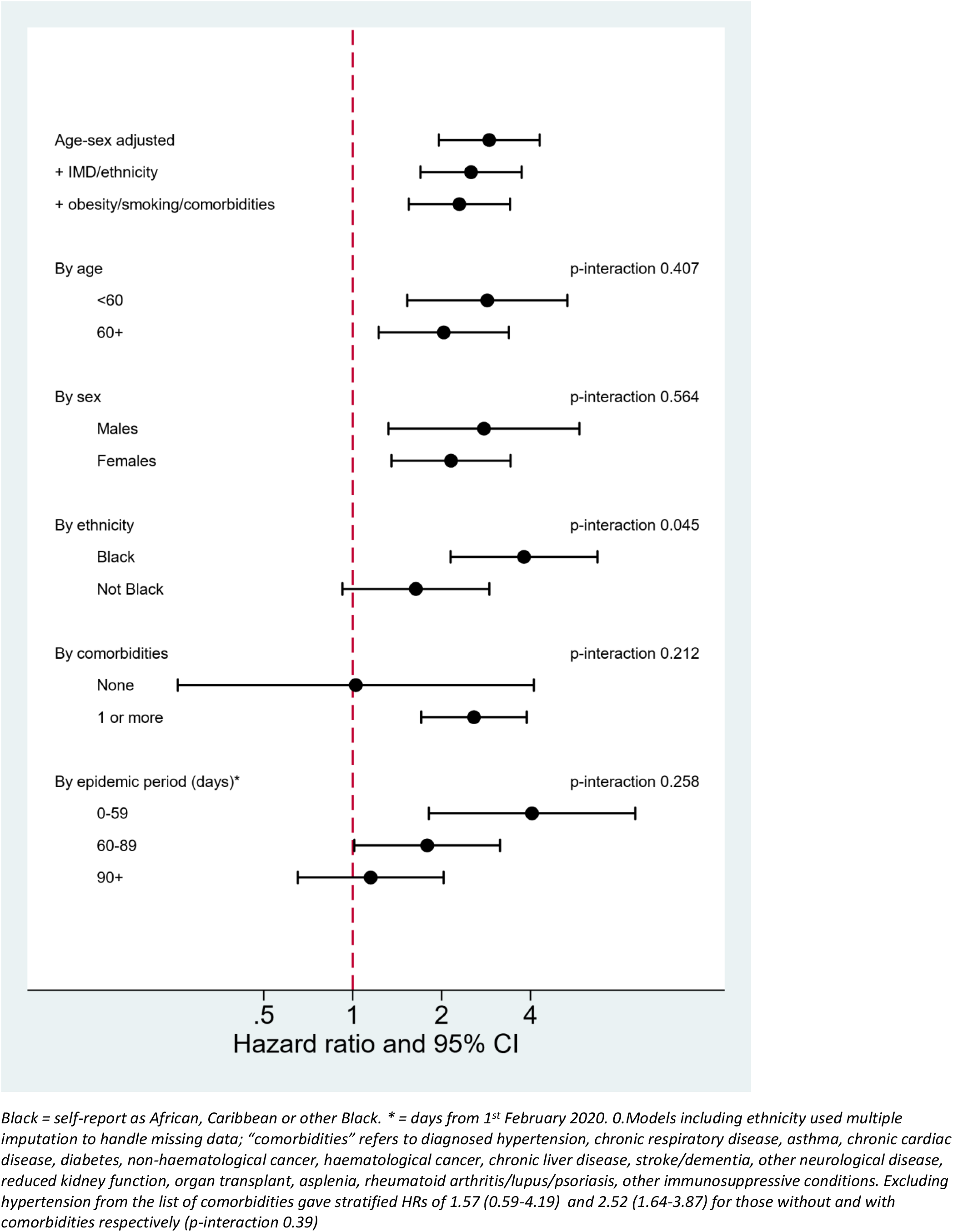
Hazard ratios for the association between HIV and COVID-19 mortality.

There was some evidence that the association between HIV and COVID-19 death was larger in black individuals (HR=3.80, 2.15-6.74, compared with 1.64, 0.92-2.90 in other ethnic groups, p-interaction=0.045, Table 3 and Figure 3). There was no statistical evidence of interaction by age, sex, or presence of other comorbidities; the point estimate for the HIV-mortality association was larger for those with at least one comorbidity but confidence intervals were wide, and excluding hypertension from the list of comorbidities attenuated the difference. The estimated association between HIV and COVID-19 death appeared to be larger early in the epidemic and reduced over time, suggesting possible non-proportionality of hazards, but this was not statistically significant (p=0.26).

Further assessment for non-proportional hazards based on Schoenfeld residuals showed no evidence of non-proportionality by HIV status (p=0.32) but there was evidence of non-proportional hazards in several adjustment covariates, namely ethnicity, sex, obesity, smoking, IMD, chronic respiratory disease, diabetes, chronic liver disease, stroke/dementia, other neurological conditions, reduced kidney function, so an additional sensitivity analysis was done modelling this non-proportionality by adding interaction terms between each of these variables and time-updated calendar time; this had very little effect on the hazard ratio for HIV (adjusted HR=2.20, 1.41-3.42 after adding interaction terms).

Results were similar in sensitivity analyses restricting to those with known ethnicity (fully adjusted HR for HIV = 2.21, 1.42-3.44), and those with complete BMI and smoking data (fully adjusted HR 1.93, 1.23-3.04).

## Discussion

### Key findings

In this large population-based study using data from the Open SAFELY platform in England, we found people with HIV to be at more than twice the risk of COVID-19 death compared to people without HIV, after accounting for demographic characteristics, lifestyle-related factors and a wide range of comorbidities. The association appeared to be particularly pronounced among people of black ethnicity, with HIV associated with a 3.8-fold higher risk of COVID-19 death in this group.

### Comparison with other evidence

Few comparable data have been published. Our results are similar to those from a population-based cohort study from the Western Cape in South Africa involving 3.5 million individuals, published as a preprint, which found an overall adjusted HR of 2.14 (1.98-3.43), compared with 2.30 (1.55-3.41) in the present study.^8^ It is noteworthy that the association was similar in an English setting with higher levels of antiretroviral treatment and viral suppression; however, this is consistent with stratified analyses in the South African data, which found a more than doubled risk of COVID-19 death even among those with a recent prescription for antiretroviral treatment and a recent viral load measure of <1000 copies/ml. Early data from small cohorts and case series in other countries included a high proportion of patients experiencing severe disease.^22-24^ A single-centre cohort in Madrid found that among 51 people living with HIV and diagnosed with COVID-19, 28 required hospital admission, and 13 had severe disease; the mortality rate among COVID-19 cases co-infected with HIV in this study was noted to be around double that in similar aged individuals in the general population.^9^ Associations between HIV and adverse COVID-19 outcomes appear to be smaller among patients already hospitalised with COVID-19: the HR for mortality was attenuated to 1.45 (1.14-1.84) in the Western Cape study when restricted to hospitalised patients,^8^ while an analysis of patients hospitalised in New York found no difference in adverse outcomes between HIV infected and demographically similar uninfected individuals.^11^ However, any role that HIV may have in increasing the risk of infection and/or development of severe disease is effectively conditioned out by restriction to hospitalised individuals who are already infected with SARS-COV-2 and likely to be experiencing severe disease at the point of inclusion.^25^

The larger association between HIV and COVID-19 death among people of black ethnicity has not previously been described. We have previously shown a larger overall risk of COVID-19 death in black and minority ethnic (BAME) compared with white groups in England,^2^ and a systematic review suggested that BAME individuals may have a higher risk of both acquiring COVID-19 infection, and experiencing worse clinical outcomes.^26^ HIV infection could plausibly exacerbate both of these, though It is not possible to delineate from our study which parts of the pathway are more or less affected by HIV status. BAME individuals are also at higher risk of negative HIV outcomes including virological rebound.^27^ Over a quarter of people with HIV in our study were black, compared to just 1.9% in the general population comparison group; understanding the reasons for the disproportionately large association between HIV and COVID-19 mortality in this group will be a priority if effective policies are to be developed to mitigate any increased risks.

### Strengths and limitations

Our study was large, so that even in a setting with a relatively low prevalence of HIV, we were able to investigate the role of HIV accounting for demographic characteristics, lifestyle-related factors including BMI and smoking, and relevant comorbidities. We tested the robustness of our findings to different missing data approaches and to non-proportional hazards. We investigated effect modification by key covariates, and were able to detect differences by ethnicity. However the relatively small number of deaths among those with HIV, reflecting the young age distribution of the HIV group, prevented definitive conclusions about the role of comorbidities, and changes in the role of HIV over time. We noted a suggestion of a larger association between HIV and COVID-19 death early in the epidemic. If confirmed, this might suggest a higher risk of infection before widespread social distancing was introduced, which could have reduced if people living with HIV were highly adherent to social distancing guidance; furthermore, some people living with HIV initially received incorrect government advice that they were considered in the highest risk category and should take extra “shielding” precautions, which might have further reduced any HIV-associated risk.^28^ We could not rule out that the observed trend over time reflected chance variation; analysis of pre-mortality indicators of severe disease, such as hospitalisation, would have greater statistical power to clarify this, but we could not obtain linked hospital data for people living with HIV, because HIV codes are considered sensitive in NHS data and transfer is legally restricted.^12,29^

For similar reasons, we were unable to include data on antiretroviral therapy use, level of viral suppression, or CD4 count, so it was not possible to stratify results, or quantify how much of the raised risk was driven by the minority of individuals with poorly controlled HIV. HIV in the UK is commonly managed in specialist clinics, but restrictions on the sharing of HIV-related codes along with policy guidance that cautions against sharing HIV-related information with the patient’s primary care provider meant that we could not access key data from routine HIV care. Whilst stratification by such factors would have added additional insights, we do not consider the lack of these data to critically undermine the utility of our findings, given that 94% of diagnosed HIV-positive individuals in the UK are on antiretrovirals and have good viral suppression (97% on therapy, and 97% of these with undetectable viral load).^13^ This high level of viral suppression in the population means that it is unlikely that the association between HIV and COVID-19 death could have been driven entirely by the 6% assumed untreated or with detectable viral load: if all deaths had been within this group, it would amount to over 1.5% becoming infected and dying from COVID-19, compared with just <0.01% of the general population. There are no available routinely-collected data on injection drug use, which could act as a confounder, though a post-hoc adjustment for history of hepatitis C infection, as a marker of possible prior injection drug use, made little difference to our results. No data are available on occupation, or any markers for contact patterns, which could have helped to evaluate the role of exposure to infection. A further limitation is that we relied on primary care records to capture HIV status. HIV diagnoses recorded in primary care are expected to be highly specific, but some underascertainment is possible; this should have had little impact on our effect estimates because HIV-positive people misclassified as HIV-negative would have made up only a tiny proportion of our comparator group. Any resulting bias is likely to have been towards the null, particularly if higher-risk people not engaged with primary care were more likely to be missed. Our outcome definition could also have been affected by some misclassification. We included clinically suspected (non-laboratory confirmed) COVID-19 deaths: due to limited testing early in the pandemic, some non-COVID-19 deaths may have been misclassified as COVID-19 deaths. We assume any such misclassification will have been non-differential with respect to HIV status, which could again have led to a degree of bias towards the null. Our study population, which was derived from one particular EHR software system, is not fully geographically representative of the broader population of England, due to substantial geographic variation in primary care provider choice of EHR software. In particular, London, which has a relatively high prevalence of HIV nationally, is under-represented in our data.

### Implications for public health

Our findings suggest that people living with HIV may be a high risk group for COVID-19-death. This indicates a need to consider targeted policies for this group as universal restrictions are eased and revised social distancing and workplace policies are introduced. People living with HIV may also need priority consideration if and when a vaccine against SARS-CoV-2 becomes available. Our findings might also have significant implications globally. Levels of antiretroviral therapy use and viral suppression in England are high, while prevalence of HIV is relatively low in global terms;^13^ the impact of HIV on the progression of the pandemic in other settings will need to be carefully monitored. Future studies should prioritise better understanding the drivers of increased COVID-19 mortality among people living with HIV, the role of ethnicity, and how the association between HIV and severe COVID-19 outcomes is affected by comorbidity status, viral load, CD4 count, and antiretroviral treatment. As well as their role in controlling HIV infection, there is also interest in whether specific antiretroviral drugs might have potential in the treatment or prevention of SARS-Cov-2 infection.^30^ In England, despite a strong overall infrastructure of linkable electronic health records data, research on these and other questions that might benefit people living with HIV is hampered by policy guidance that has generally led to restrictions in the sharing and flow of HIV-related data, despite apparent support for sharing for public health purposes.^31^ We propose a review of guidance to address this and ensure that people living with HIV are not unnecessarily excluded from important research.”

### Conclusion

In this large population-based study, people living with HIV in England had more than double the risk of COVID-19 death than people without HIV, after accounting for demographic characteristics, lifestyle-related factors and comorbidities. Monitoring the role of HIV on COVID-19 outcomes in other settings will be crucial as the pandemic progresses.

## Data Availability

All data were linked, stored and analysed securely within the OpenSAFELY platform https://opensafely.org/. All code is shared openly for review and re-use under MIT open license. Detailed pseudonymised patient data is potentially re-identifiable and therefore not shared. We rapidly delivered the OpenSAFELY data analysis platform without prior funding to deliver timely analyses on urgent research questions in the context of the global Covid-19 health emergency: now that the platform is established we are developing a formal process for external users to request access in collaboration with NHS England; details of this process will be published shortly on OpenSAFELY.org.

## Author contributions

BG conceived the OpenSAFELY platform and the approach. BG and LS lead the project overall and are guarantors. KB designed the study, did the analysis and wrote the first draft. SB led on software development. AM led on information governance. Other contributions were – data curation – CB, JP, JC, SH, SB, DE, PI and CEM; disease category conceptualisation and codelists – KB CTR BM CB JC CEM AJW HIM HJF HJC JP; statistical analysis code: KB, EW; ethical approvals – HJC EW LS BG; software – SB, DE, PI, AJW, WH, CEM, CB, FH, JC; writing (reviewing and editing) – KB, CTR, BM, AS, AM, RME, CEM, IJD, SJWE, LS, BG. All authors were involved in design and conceptual development and reviewed and approved the final manuscript.

## Data sharing

All data were linked, stored and analysed securely within the OpenSAFELY platform https://opensafely.org/. All code is shared openly for review and re-use under MIT open license (https://github.com/opensafely/hiv-research). Detailed pseudonymised patient data is potentially re-identifiable and therefore not shared. We rapidly delivered the OpenSAFELY data analysis platform without prior funding to deliver timely analyses on urgent research questions in the context of the global Covid-19 health emergency: now that the platform is established we are developing a formal process for external users to request access in collaboration with NHS England; details of this process will be published shortly on OpenSAFELY.org.

